# Psychosocial factors associated with mental health and quality of life during the COVID-19 pandemic among low-income urban dwellers in Peninsular Malaysia

**DOI:** 10.1101/2022.02.21.22271310

**Authors:** Wong Min Fui, Hazreen Abdul Majid, Rozmi Ismail, Tin Tin Su, Tan Maw Pin, Mas Ayu Said

## Abstract

**Background and aims:** The mental well-being among low-income urban populations is arguably challenged more than any other population amid the COVID-19 pandemic. This study investigates factors associated with depression and anxiety symptoms, and quality of life among Malaysia’s multi-ethnic urban lower-income communities.

**Methods:** This is a community-based house-to-house survey conducted from September to November 2020 at the Petaling district in Selangor, Malaysia. Five hundred and four households were identified using random sampling, and heads of eligible households were recruited. Inclusion criteria were age ≥ 18 years with monthly household income ≤RM6960 (estimated USD 1600) without acute psychiatric illness. The PHQ-9, GAD-7 and EQ-5D were used for depression, anxiety, and quality of life. Multivariable logistic regression was performed for the final analysis.

**Results:** A total of 432 (85.7%) respondents with a mean age of 43.1 years completed the survey. Mild to severe depression was detected in 29.6%, mild to severe anxiety in 14.7%, and problematic quality of life in 27.8% of respondents. Factors associated with mild to severe depression were younger age, chronic health conditions, past stressful events, lack of communication gadgets and lack of assets or commercial property. While respiratory diseases, marital status, workplace issues, financial constraints, absence of investments, substance use and lack of rental income were associated with mild to severe anxiety. Not attributing poverty to structural issues, help-seeking from professionals, and self-stigma were barriers, while resiliency facilitated good psychological health. Problematic quality of life was associated with depression, older age, unemployment, cash shortage, hypertension, diabetes, stressful life events and low health literacy.

**Conclusions:** A higher proportion had reported mild to severe anxiety and depression symptoms in the sampled urban poor population than previous pre-pandemic reports. The psychosocial determinants should inform policy and direct future research within this underserved population.

## Introduction

Depression and anxiety remain two major diagnosable common mental disorders contributing to disability and morbidity worldwide, partially attributable to lack of public health investment in this area (1). The World Health Organisation reported 800,000 suicides cases globally in 2015, with 78% of suicides occurring in the lower to middle-income countries (LMIC) (2). Almost half a million Malaysian have significant depressive symptoms that are particularly prominent among individuals living within households within the bottom 40% (B40) income bracket (3). The recent coronavirus disease (COVID-19) pandemic has brought mental health issues to the forefront, with a reported increase in suicide cases from 609 cases in 2019 to 631 cases in 2020 during the pandemic period (4,5).

Over the past 50 years, the Malaysian government has invested in extreme poverty eradication and economic growth by shared prosperity within its multi-ethnic population. The rapid urbanisation observed from 1960 to 2010 (6) had, however, led to the unintended consequences of the transformation of social structures resulting in pockets of urban poor in the cities of Malaysia. This has led to escalating poverty in urban areas within Selangor, the wealthiest state in Peninsular Malaysia (7), which threatens urban residents’ mental well-being (8). Studies from high-income countries (HIC) and low-middle-income countries (LMICs) suggest that urban residents were more likely to develop neurotic conditions than rural residents (9-11). In contrast, studies from China and Germany showed that rural residents were prone to mental health issues (12,13). Local studies also revealed a consistently high prevalence of depression (12.3% to 24.2%) and anxiety (18% to 36.3%) among the low-income urban residents, with variations arising from on screening tools utilised and selected cut-off scores (14-16).

Low socioeconomic status is a major social determinant of health (SDH) (17), which profoundly affects the morbidity and mortality of the community (18). Education, ethnic group or social class, income, and employment are typical indicators for socioeconomic status (19). Community-based studies from LMIC have revealed the relationship between low socioeconomic status (SES) and common mental disorders (20-23).

Recent literature revealed that low socioeconomic status was linked to lower health literacy with higher stigma (24,25) and lack of mental health help-seeking (26). Additionally, low-income individuals’ perceptions of the structural issues they encountered may subsequently influence their decision-making, thus influencing mental health (27). Since the evidence for these psychosocial factors remains scarce, further exploration is needed to target mental health promotion among low-income populations (26).

A study conducted during the COVID-19 pandemic revealed that those who showed avoidance coping and lower religious coping had a higher risk of developing mild to moderate depressive symptoms (28). A recent study from China showed that resilience scores were inversely associated with mental health symptoms associated with lower among subjects with mild COVID-19 cases (29). This has led to concerns that the COVID-19 pandemic may adversely affect the mental health of populations in LMIC that lack the resources to address the increase in mental health needs of their population.

In addition, the World Bank has estimated that between 71 to 100 million people are being pushed into poverty due to the COVID-19 pandemic (30). Therefore, the current study measured the prevalence of depression and anxiety symptoms and quality of life among the urban low-income population in Malaysia during the COVID-19 and identified psychosocial determinants of depression, anxiety, and quality of life within this population.

## Methods

### Setting

This was a community-based cross-sectional survey conducted from September 2021 until November 2021 (corresponds to the recovery movement control order (RMCO) and start of the Controlled Movement Control Order (CMCO)) (See Fig 1) at the Petaling district of the state of Selangor in Malaysia. Proportions and effect sizes were obtained from similar studies to estimate sample size (S1 Table). The sample size was estimated using Open Epi software with a significance level of 0.05 and an alpha value of 0.2.

**Fig 1.**
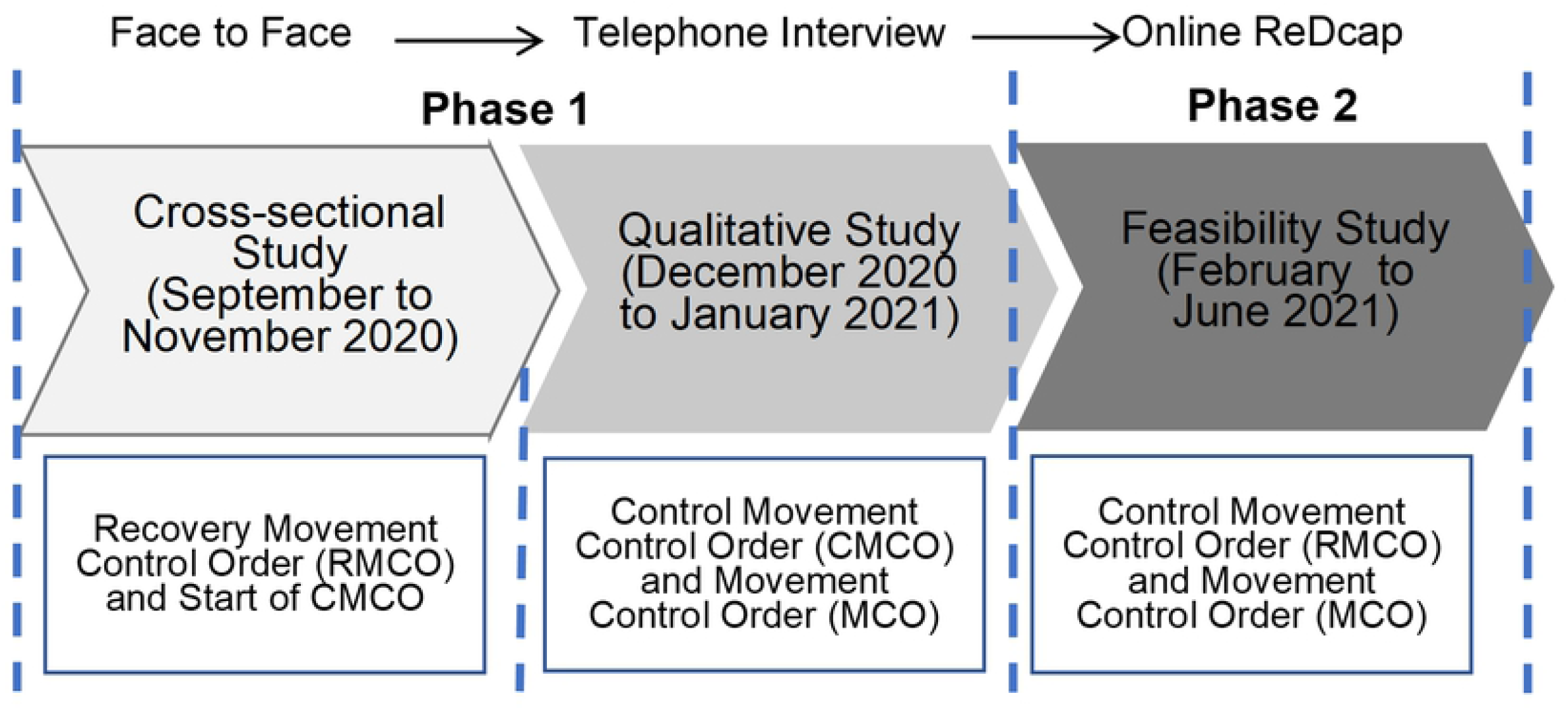
Timeline of each phase of study and the corresponding movement control.

A simple random sampling was done based on the household list provided by the research committee from the Statistics Department. All respondents from the selected household who was a) aged 18 years and older with, (b) a household income of RM 6960 and below, and c) without any acute psychiatric illness were included in this study. Non-Malaysians were excluded from the study. This study received ethical approval from the University of Malaya Research Ethics Committee (UMREC Non-Medical ref: UM.TNC2/UMREC – 811). Written consent was obtained from each eligible respondent prior to the enrolment.

### Data Collection

Since this is a face-to-face data collection, each enumerator was briefed about the University Malaya COVID-19 Fieldwork Safety Protocol. Trained enumerators administered validated questionnaires during the house-to-house data collection(See Fig 2).

**Fig 2.**
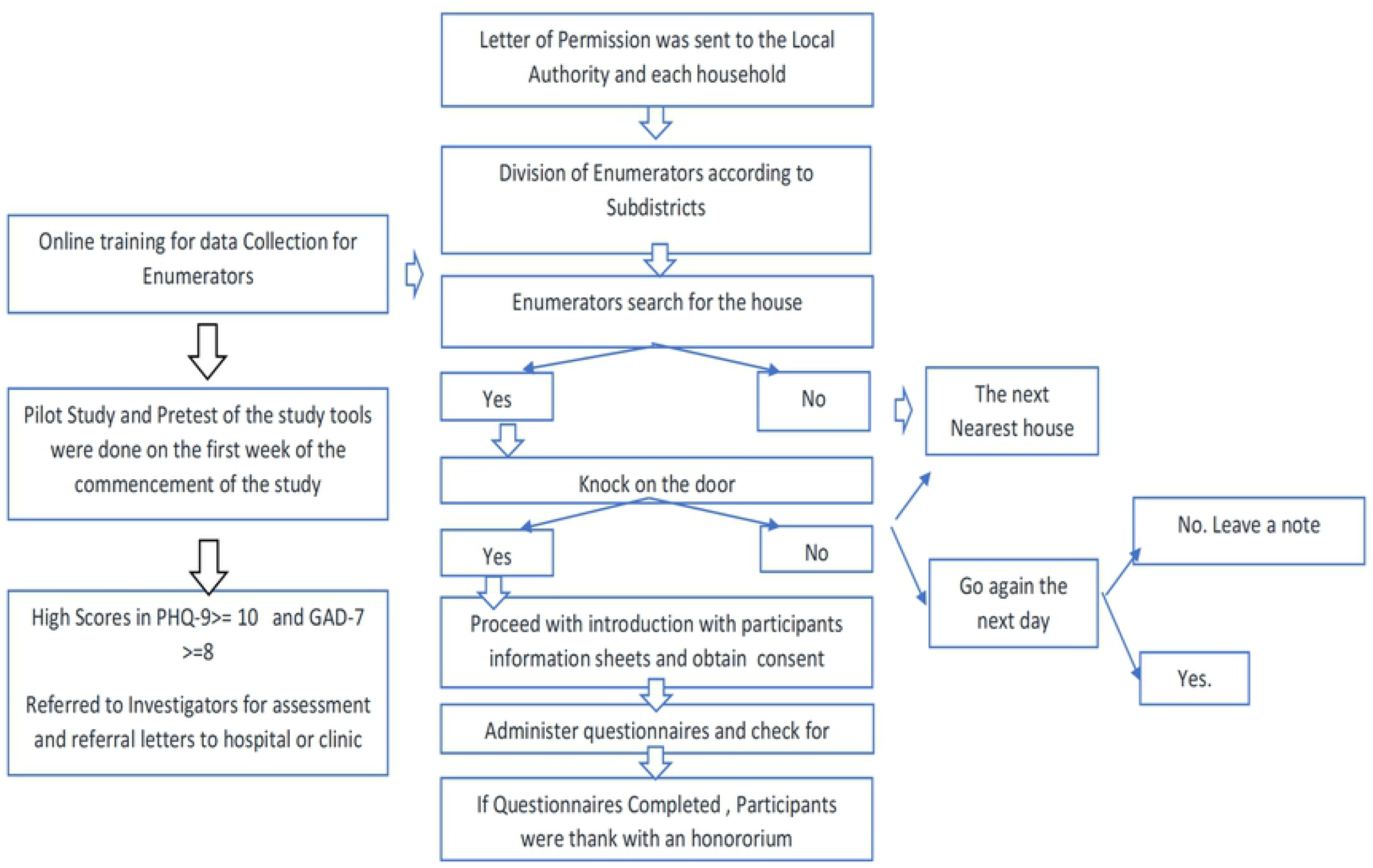
The flow chart of the data collection.

The respondents completed multiple standardised instruments in the Malay language with assistance from the research team. The survey components comprised the socio-demographic domain which captured the respondents’ information such as age, gender, ethnicity, education level, employment status during the COVID-19 pandemic, monthly household income during COVID-19 pandemic, marital status and household size.

The health domain components comprised self-reported weight and height, history of chronic illnesses, stressful life events, substance used, the 6-item health literacy scale (HL-6) (Duong et al., 2017), 9-item Patient’s Health Questionnaire (PHQ-9) (31), 7-item Generalised Anxiety Disorders (GAD-7) (32) and the EQ-5D-5L health-related quality of life (33). All the study tools were piloted before the actual study, and all of them showed good internal consistency (refer to Table S2).

The presence of chronic diseases was self-reported by the participants based on the diagnoses from medical professionals and was defined as conditions that lasted for one year or more that require ongoing medical attention or limiting daily activities, or both. The listed medical conditions included hypertension, diabetes mellitus, heart diseases, stroke, mental illness, and cancer. Substance use was recorded with a checklist of the top 10 most common substances, including tobacco, alcohol, cannabis, cocaine, amphetamine-type stimulant, inhalant, sleeping pills, hallucinogens, opioids and other substances. The 13-item Malay version of the stressful life events checklist was also utilised (34).

EQ-5D-5L is a standardised self-reported perceived health-related quality of life (QoL) which is rated through a descriptive system (EQ-5D) and a visual analogue scale (EQ-VAS), which captures perceptions on health status in the mobility, selfcare, usual activities, pain or discomfort, and anxiety or depression domain. Each dimension is rated based on five response levels from no problems to unable to/extreme problems. Each health state is assigned a summary index score derived from a country-specific value set (35). Health state index scores ranged from less than 0 to 1 (the value of full health), with higher scores indicating higher health utility. The visual analogue scale (EQ-VAS) records the respondent’s overall current health on a vertical visual analogue scale on a scale of 0 to 100. The Cronbach’s alpha of the EQ-5D was 0.85 (36).

The Patient Health Questionnaire measured depressive symptoms over the past two weeks. The PHQ-9 consists of 9 items scored across a four-point scale with a maximum score of 27. It is then classified further according to minimal (0-4), mild (5-9), moderate (10-14), severe (15-19), and very severe (20 or greater). Both English and Malay versions of the questionnaire has good internal reliability with a Cronbach’s α of 0.70 (31, 37). The total scoring methods were adopted (38).

Generalised Anxiety Scale also measures anxiety symptoms for the past two weeks. This was a 7-item instrument rated on a four-point scale. These cut-off values of minimal (0-4), mild (5-9), moderate (10 -15) and severe (16-21) (39) were utilised. The Malay and English versions had good internal reliability with Cronbach’s α ranging from 0.74 to 0.92 (32, 40).

Religiosity was determined with the Santa Clara Strength of Religious Faith Questionnaire 5 items (SCSRFQ-5) brief version, which was self-reported and rated on a 4-point Likert scale with a maximal score of 20 and a higher score indicating greater strength religiosity. The cut-off point for high (≥ 17) or low (<17) religiosity were based on the sample median (41). This tool was validated in the pilot study with good internal reliability of Cronbach’s α of 0.79.

Resilience Scale 14 items (RS-14) was derived from the 25 items scale developed by Wagnild et al. (42). Each item was rated with a 7-point scale with a maximum total score of 98. The Malay version of RS-14 has an excellent internal consistency of 0.86 (43).

Mental Help-Seeking Attitudes Scale (MHSAS) is a 9-item semantic scale with a higher score indicating a more positive attitude towards seeking help (44). The Malay MHSAS has a Cronbach’s α of 0.892 (45).

Self-Stigma of Seeking Help (SSOSH) scale was developed by Vogel et al. and has 10 items rated on a 5-point scale. The Malay SSOSH has been validated among the low-income group in Malaysia with an acceptable internal consistency of Cronbach’s alpha = 0.667 (26).

Poverty Attribution was used to measure the perception of low-income respondents on the cause of poverty. This 21-item new tool was developed by Rozmi et al. (46) and comprised structural support, socioeconomic support, individualistic and fatalistic domains. Each item is rated on a 5-point scale with higher scores indicating higher levels of agreement on the cause of impoverished conditions. The Malay language version showed acceptable internal validity of Cronbach alpha 0.61 to 0.87 among the four main domains.

Health Literacy Survey was a valid and reliable (Cronbach’s alpha=0.85) tool in six Asian countries (47). Six items were extracted from the 12-item short-form version, with each item rated on a 4-point Likert scale. This tool was validated in the pilot study with good internal consistency (Cronbach’s alpha=0.898) and test-retest of inter-item covariance of 0.469 with excellent Pearson correlation of r (98) = .839, p< .0001 were obtained. The total scores were computed and utilised for data analysis.

## Statistical Analysis

### Data Analysis

All statistical analyses were performed using the IBM SPSS Statistics for Windows, version 25.0 (IBM Corp LP, Armonk, NY, USA). Total scores of ≥ 5 for PHQ-9 and GAD-7 was applied to determine the presence of symptomatic depression (depressed =1, not-depressed =0) and positive symptomatic anxiety (anxious=1, not-anxious =0), respectively. The students’ independent sample T-test (normally distributed variables) and Mann-Whitney U-test (non-normally distributed variables) were conducted for univariate analysis. The Pearson’s Chi-square or Fisher’s Exact test was used to determine the strength of association between categorical explanatory variables and the outcomes variables. Multivariable logistic regression was performed to elicit significant final independent variables associated with PHQ-9, GAD-7 and EQ-5D. Hosmer-Lemeshow model development strategy was applied in the final analysis with variables selection criterion of p-values of less than 0.25 with backwards purposive variables exclusion of p-value more than 0.05 guided by the likelihood-ratio test (48). Effect modification first-order main effects of explanatory variables were checked guided by the likelihood-ratio test. The finalised model was assessed for violation of assumptions of linearity of explanatory variables, log odds, multicollinearity and model fitness. The model’s sensitivity was also assessed using the area under receiving operating characteristics (AUROC) curve. Statistical significance was set at a p-value less than 0.05.

## Results

### Participant Characteristics

A total of 504 eligible participants, 432 (82.7%) participants completed the survey. Table 1 summarises participants’ characteristics. Respondents had a mean age of 43.1 (SD 13.2) years. The unemployment rate doubled (33.2%) during the pandemic while income reduced by 13.5 %, and 41% lived below the poverty line.

**Table 1.** Socio-demographic profiles of Low-income respondents from the Petaling district (n=432)

| Variables | n | % |
| --- | --- | --- |
| **Gender** |  |  |
| Male | 278 | 64.5 |
| Female | 153 | 35.5 |
| **Age group (years old)** |  |  |
| Below 30 | 81 | 19.8 |
| 30-40 | 114 | 27.8 |
| 41-50 | 104 | 25.4 |
| More than 50 | 111 | 27.0 |
| **Ethnicity** |  |  |
| Malay | 323 | 75.1 |
| Chinese | 58 | 13.5 |
| Indian | 31 | 7.2 |
| Others | 18 | 4.2 |
| **Marital status** |  |  |
| Single | 75 | 17.5 |
| Married | 321 | 74.7 |
| Widowed | 34 | 7.8 |
| **Education level categorical** |  |  |
| No Formal & Primary School | 29 | 6.8 |
| Secondary Schools | 206 | (48.4) |
| More than Secondary Schools | 191 | (44.8) |
| **Employment status (before Covid-19)** |  |  |
| No | 69 | 16.2 |
| Yes | 357 | 83.8 |
| **Employment status (during Covid-19)** |  |  |
| Yes | 276 | 66.8 |
| No | 137 | 33.2 |
| **Household income (during Covid-19)** |  |  |
| Below MYR 1700 | 136 | 31.5 |
| MYR 1700 to 2700 | 91 | 21.1 |
| MYR 2701 to 3700 | 77 | 17.8 |
| MYR 3701 to 4700 | 55 | 12.7 |
| MYR 4700-5700 | 32 | 7.4 |
| More than MYR 5701 | 41 | 9.5 |
| **Household size (person)** |  |  |
| Less than 4 | 191 | 47.8 |
| More than 4 | 234 | 55.2 |
| **House-ownership** |  |  |
| Owners | 179 | 41.7 |
| Shelter | 13 | 3.0 |
| Renting | 219 | 51.1 |
| Inherited | 18 | 4.2 |
| **Asset** |  |  |
| House | 231 | 53.8 |
| Vehicle | 350 | 81.8 |
| Land | 42 | 9.8 |
| Orchard | 23 | 5.4 |
| Cash | 148 | 34.5 |
| Rental house | 55 | 12.9 |
| Jewellery | 128 | 29.9 |
| Shophouse | 6 | 1.4 |
| Investment | 49 | 11.4 |
| Others | 4 | 0.9 |
| **Communication tools (own a mobile or smartphone)** |  |  |
| Yes | 340 | 81.9 |
| No | 75 | 18.1 |
| **Own at least a form of communication tools (TV or radio and internet or laptop and smartphone/mobile phone)** |  |  |
| Yes | 405 | 97.59 |
| No | 10 | 2.41 |

Approximately 133 (31.2%) respondents had problematic literacy levels, and 29 (6.8%) had an inadequate level of health literacy. The professional help-seeking median score was high at 6.0 (IQR 2), and the mean self-stigma was neutral at 3.0 (SD 0.6). Poverty attribution, structural and socioeconomic domains scores were 3.6 (SD 0.8) and 3.8 (SD 0.9), respectively. Low resiliency was found in 133 (31%) participants. 182 (42%) scored 17 and below for religiosity. Of the total respondents, 432 (30.4%) had a self-reported history of non-communicable diseases with hypertension in 78 (18.2%), diabetes 38 (8.9%) and respiratory disease (COPD or Asthma) 20 (4.7%). 172 (40.7 %) were overweight (body mass index (BMI) >23.0-27.4 kg/m^2^) and 29.6 % obese (BMI ≥27.5kg/m^2^). In 130 (30.2%) male respondents, substance use was present, with tobacco, alcohol, and sleeping pills being the commonest used substances. Approximately 209 (48.5%) had a history of stressful life events, including losing loved ones, followed by working environment issues and job loss.

Mild to severe depression symptoms were reported by 127 (29.6 %), 30 (7%) (n =30) of whom had moderate to severe depressive symptoms. Mild to severe anxiety symptoms were present in 63 (14.6%), with 19 (4.4%) reporting moderate to severe depressive symptoms. For the details of the descriptive findings, refer to Tables S3 and S4.

### Multivariable Analyses

Variables positively associated with mild to severe symptoms of depression were those aged less than 30 years OR 5.11 (95%CI 2.04, 12.83), self-reported hypertension, having other chronic illnesses, presence of past stressful life events (physical assault, long term illness, family issues and workplace issues). Protective factors against the development of mild to severe depressive symptoms were those who owned one or more communication tools (Television or radio and internet or laptop and smartphone/mobile), absence of assets such as investment shares, shop-houses, attributing structural issues to poverty, increase in resilience scores and those who finds professional help is beneficial. Factors associated with anxiety symptoms were respiratory illness (COPD or Asthma), stressful life events (marital issues, financial issues, workplace issues), and sleeping pills. Perceived structural issues to poverty, less self-stigma, and higher resilience scores were associated with fewer anxiety symptoms (Table 2). Quality of Life (QoL) was problematic in 119 (27.8%) respondents. Mild to severe depressive symptoms were associated with increased odds of problematic QoL. Age, female sex, unemployment, hypertension, diabetes, stressful life events of severe injury were associated with poorer QoL. Higher literacy scores were protective against poorer QoL (Table 3). Refer to Table S5 A-I for the details of univariable analysis and Table J-L for the multivariable variable selection process.

**Table 2:** Simple and multivariable logistic regression on factors associated with Depression (n=385) and Anxiety Symptoms(n=398) (cut-off at 5)

| Factors | Depression (PHQ-9) |  | Anxiety (GAD-7) |  |
| --- | --- | --- | --- | --- |
|  | Crude OR (95% CI) | Adjusted OR (95% CI) | Crude OR (95% CI) | Adjusted OR (95% CI) |
| **Age** |  |  |  |  |
| Below 30 | Reference | Reference | Reference | Reference |
| 30-40 | 0.45 (0.24, 0.83)* | 0.52 (0.24, 1.12) | 0.68 (0.32, 1.45) | 0.78 (0.31, 1.92) |
| 41-50 | 0.46 (0.24, 0.88)* | 0.47 (0.21, 1.09) | 0.51 (0.22, 1.17) | 0.68 (0.25, 1.88) |
| More than 50 | 0.35 (0.18, 0.67)* | 0.20 (0.08, 0.49)* | 0.47 (0.20, 1.07) | 0.41(0.15, 1.12) |
| **Asset-Shop lot** |  |  |  |  |
| Yes | Reference | Reference |  |  |
| No | 0.08 (0.01, 0.70)* | 0.08 (0.07, 1.00) |  |  |
| **Asset-Investment** |  |  |  |  |
| Yes | Reference | Reference |  |  |
| No | 0.52 (0.28, 0.95)* | 0.37 (0.17, 0.80)* |  |  |
| **Asset-Rental house** |  |  |  |  |
| Yes |  |  | Reference | Reference |
| No |  |  | 0.65 (0.32, 1.34) | 0.44 (0.18, 1.06) |
| **Hypertension** |  |  |  |  |
| No | Reference | Reference |  |  |
| Yes | 1.57 (0.94, 2.63) | 2.25 (1.01, 5.00)* |  |  |
| **Other diseases** |  |  |  |  |
| No | Reference | Reference |  |  |
| Yes | 7.29 (2.78, 19.10)** | 4.06 (1.26, 13.06)* |  |  |
| **Asthma** |  |  |  |  |
| No |  |  | Reference | Reference |
| Yes |  |  | 2.12 (0.74, 6.07) | 4.44 (1.18, 16.67)* |
| **Sleep pill usage** |  |  | Reference | Reference |
| No |  |  | Reference | Reference |
| Yes |  |  | 10.46 (2.43, 44.95)* | 11.89 (1.26, 112.16)* |
| **Physical or sexual assault** |  |  |  |  |
| No | Reference | Reference |  |  |
| Yes | 5.68 (2.38, 13.54)** | 4.77 (1.56, 14.57)* |  |  |
| **Prolonged or serious illness** |  |  |  |  |
| No | Reference | Reference |  |  |
| Yes | 5.24 (2.45, 11.236)** | 4.892 (1.834, 13.053)* |  |  |
| **Family Issues** |  |  |  |  |
| No | Reference | Reference |  |  |
| Yes | 6.87 (2.94, 16.06)** | 3.74 (1.26, 11.14)* |  |  |
| **Marital Problem** |  |  |  |  |
| No |  |  | Reference | Reference |
| Yes |  |  | 8.40 (3.45, 20.44)** | 4.73 (1.52, 14.70)* |
| **Financial issues** |  |  |  |  |
| No |  |  | Reference | Reference |
| Yes |  |  | 8.45 (3.70, 19.32)** | 4.67 (1.46, 14.93)* |
| **Workplace Issue** |  |  |  |  |
| No | Reference | Reference | Reference | Reference |
| Yes | 3.28 (1.78, 6.04)** | 3.48 (1.58, 7.67)* | 5.03 (2.61, 9.71)** | 3.533 (1.53, 8.17)* |
| **Owing Communication Gadget 1 or 2** |  |  |  |  |
| **No** | Reference | Reference |  |  |
| Yes | 0.27(0.07, 0.97)* | 0.06 (0.01, 0.38)* |  |  |
| **Poverty attribute-Structural** | 0.65 (0.51, 0.83)** | 0.66 (0.48, 0.91)* | 0.78 (0.58, 1.05) | 0.67 (0.45, 0.99)* |
| **MHSAS (Total)** | 0.96 (0.94, 0.98)** | 0.96 (0.94, 0.99)* | 0.98 (0.95, 1.00) | 0.99 (0.97, 1.03) |
| **SSOSH (Total)** |  |  | 1.12 (1.06, 1.19)** | 1.09 (1.01, 1.17)* |
| **Resilience (Total)** | 0.96 (0.94, 0.97)** | 0.96 (0.94, 0.98)** | 0.96 (0.94, 0.98)** | 0.97 (0.95, 0.998)* |

OR = Odds ratio; CI = Confidence Interval. \*\* p = < 0.001 \* p=<0.05
PHQ-9: Nagelkerke's R square = 0.372; Hosmer and Lemeshow Test = 0.382; Area under the curve = 0.837 p = < 0.001
GAD-7: Nagelkerke's R square = 0.307; Hosmer-Lemeshow goodness of fit test = 0.888., Area under the curve = 0.799 (95% CI: 0.737, 0.861, p = < 0.001)

**Table 3:** Simple and multivariable logistic regression on factors associated with EQ-5D (n=385)

| Factors | Crude OR (95% CI) | Adjusted OR (95% CI) |
| --- | --- | --- |
| **Gender** |  |  |
| Male | Reference | Reference |
| Female | 1.61 (1.05, 2.49)* | 2.34(1.35, 4.04)* |
| **Age group (year-old)** |  |  |
| Below 30 | Reference | Reference |
| 30-40 | 1.09 (0.51,2.34) | 1.21 (0.49, 2.99) |
| 41-50 | 1.53 (0.71, 3.28) | 1.61(0.63, 4.10) |
| More than 50 | 3.24 (1.56, 6.74)* | 3.06 (1.17, 8.03)* |
| **Asset-Cash** |  |  |
| Yes | Reference | Reference |
| No | 1.61(1.01, 2.56)* | 1.88 (1.04, 3.39)* |
| **Work during outbreak** |  |  |
| Yes (R) | Reference | Reference |
| No | 2.33(1.49, 3.64)** | 1.34 (0.76, 2.38) |
| **Hypertension** |  |  |
| No | Reference | Reference |
| Yes | 4.18 (2.49, 6.99)** | 2.56 (1.22, 5.26)* |
| **Diabetes** |  |  |
| No | Reference | Reference |
| Yes | 5.98 (2.93, 12.22)** | 3.07 (1.22, 7.71)* |
| **Severe injury due to accident** |  |  |
| No | Reference | Reference |
| Yes | 2.88 (1.41, 5.86)* | 3.59 (1.37, 9.42)* |
| **Jobless** |  |  |
| No | Reference | Reference |
| Yes | 2.47 (1.35, 4.59)* | 2.54 (1.20, 5.37)* |
| **Depression-symptomatic** |  |  |
| < 5 (R) | Reference | Reference |
| ≥ 5 | 3.31 (2.11, 5.18)** | 2.79 (1.57, 4.49)** |
| **Health literacy index** | 0.96 (0.94 , 0.98)** | 0.96 (0.94, 0.99)* |

OR = Odds ratio; CI = Confidence Interval. \*\* p = < 0.001 \* p=<0.05
Nagelkerke's R square = 0.345; Hosmer and Lemeshow Test = 0.857; Area under the curve = 0.813 (95% CI: 0.765, 0.861), p = < 0.001

## Discussions

Comparatively, depression and anxiety levels were higher than those previously reported in the National Health and Morbidity Survey (NHMS) 2012 -2019(3,49). Compared to other local studies, anxiety levels were lower than the pandemic levels (28,50-51). While mild to moderate anxiety levels were higher than those previously reported in international studies (52-54), it was also lower than those conducted in other countries during the pandemic (55, 56). Lower-income groups with higher financial strain respondents are more likely to experience mild to severe depression (28). However, the lower anxiety levels could be attributed to differences in assessment tools (28,50-51). As the study was conducted towards the later stages of the pandemic as opposed to previous studies, (57,58) anxiety levels could have been higher initially due to difficulties adapting to containment measures and fear of infection and death (59). Depression may be more prominent at the later stages when the future is uncertain due to job losses, pay cuts and poor social support (60,61).

To date, only a few studies have examined the relationship between mental health issues and quality of life for low-income groups that allowed for comparison. The majority were those studies targeted at the general population with specific diseases (62). Nevertheless, the comparisons were made based on studies that involved the general population with a similar methodology. The overall prevalence for poorer quality of life was 27.8%, which is lower than the majority of the pre-pandemic studies from other countries (63-67) and remained low even after accounting for socio-demographic and chronic illness characteristics (64,68). Out of this figure, pain/discomfort and anxiety/depression domains had the most significant number of problematic QoL respondents. Local evidence remains fragmented and focuses primarily on the diseased population but not on the general population. The possible explanation for the lower figure is the higher proportion of Malay and male participants in this study sample. In a separate cross-tabulation analysis done in the present study, anxiety/depression was the only domain that showed significant difference across ethnic groups and gender, with Malay and males inclining to report “no problem” to their mental health well-being. Another local validation study observed a similar trend (69). Therefore, the better-perceived health-related quality of life among B40 for the current study is worth further exploration in the future study to rule out the possible information bias due to cultural influence on the lack of disclosure of mental health issues.

A positive association between higher scores of depression and poorer quality of life were established in this study. This results were similar with studies conducted in Malaysia, China and Slovenia involving urban community samples (28, 70-71). Leong et al. from the northwest coast of Peninsular Malaysia has conducted a similar study with different tools, and results varied for depression and anxiety among the domains in WHOQoL-BREF (50). Another large scale mental health study National Epidemiologic Survey on Alcohol and Related Conditions, has shown (NESARC) (72) reported that anxiety does not confer a significant association with quality of life. Variation in methodology in terms of the measurement tools and location of the studies may explain the inconsistency of findings.

The escalation of financial burdens due to pay cuts and unemployment has led to marital and family issues during the pandemic, increasing depression and anxiety symptoms. This was supported by a Malaysian study done in urban settings that revealed severe problems at work, unhappy relationships with children, spouse and family, and severe financial constraints are stressful life events for depression (73). The same investigator also measured anxiety symptoms and found that unhappy relationships with family and severe problems at work predict the outcomes (74). The odds ratio for stressful life events were two times higher than those pre-pandemic findings reported by Kader et al. (73). No doubt, movement restriction increases contact time among the family members, but it can aggravate the pre-existing family conflict and causes stress to the vulnerable population (75). In line with the current study’s findings, physical or sexual assault showed a positive relationship with depression. Woman’s Aid Organisation (WAO) and Women, Family and Community Development Ministry reported increased usage of their public hotlines during the pandemic, and domestic violence was the main reason for calling (76).

Additionally, Malaysia’s divorce rate also rises from 60,088 cases in 2017 to 90,766 cases in 2020, based on the syariah court data (77). The majority of the marital issues involved cases from the bottom 40 % of the low-income group who faced the challenge of job loss and financial crisis (78). The economic difficulty is closely related to depression among the parents (79). Therefore, it is pertinent to look out for stressful life events like a history of child abuse or intimate partner physical abuse as a critical risk factor for mental health issues among the low-income group.

Younger respondents (less than 30 years old) have a higher risk of developing depression symptoms than older respondents during the pandemic. The social, emotional and cognitive maturation was observed by neurodevelopmental scientists even right before the adolescent age extending to the mid-20s to the 30s of age (80,81); this has marked the vulnerability of the brain towards environmental changes and insults during this transition period towards young adulthood, thus any stressful life event will tip off mental health issues. Three studies were conducted in the local community, and the general population at the urban settings supported the relationship between the younger age group of respondents and a higher likelihood of mental health issues (14-15,28). Likewise, similar findings are notable among studies in other countries (82-84).

Gender did not reveal any positive findings for mental health status in this study. These were unexpected findings as females were the most replicable risk factors in the past studies among the general population (85, 86), low-income groups (21, 87), and during the pandemic (82, 87, 88) (A. Ahmad, Rahman, & Agarwal, 2020; Huang & Zhao, 2020). Perhaps the COVID-19 pandemic may have put the low-income family under serious financial strain, leading to emotional turmoil for both genders. On the other hand, the older age group (more than 50 years) and females had a poorer quality of life. The plausible explanation is the higher probability of getting chronic illness at an older age, and chronic illness tend to be higher among females than males based on the NHMS 2019 data (3).

Non-communicable diseases, including hypertension and diabetes, were reported by one in three persons within the low-income population which approximated to the prevalence of known hypertension of 15.5% and diabetes of 9.2% from the urban population reported in the NHMS 2019 (3). In contrast, a meta-analysis reported by Uphoff, Newbould (89) showed a six to seven times higher pooled prevalence than this study. The higher rate observed in the latter study was attributed to hospital-based samples and underdeveloped mental health services for secondary prevention in lower-income countries.

Respondents with a known history of hypertension, other diseases and perceived chronic illness as stressful life events likely to report higher depressive symptoms. Asthma was associated with a higher risk for anxiety symptoms. However, out of the self-reported illness, only hypertension and diabetes were associated with poorer QoL. Studies showed that physical illnesses like cardiovascular-related diseases had proven to predict common mental health issues (90) and affect the quality of life among the low-income population. There was convincing evidence from a systematic review with meta-analyses done by Kohler et al. (91). that both obesity and metabolic syndrome predict the occurrence of depression studies for anxiety disorders involved the general population have revealed that pre-morbid chronic conditions predict psychiatric illness (aOR 3.66 95% CI:1.62, 8.30) (92,93). The plausible explanation for chronic and mental illnesses was the disruption of their quality of life due to the complication of the chronic illness. For instance, B40 respondents who developed diabetes nephropathy and retinopathy may face significant stress due to the impairment apart from their medical financial burden.

Respondents reported a high proportion of low health literacy, with positive findings and poorer life quality. This evidence supported the high rate of chronic illness among the low-income group revealed by a series of publications from PARTNER’s (94-97). However, collective evidence from the meta-analysis showed heterogeneous results for the association between health literacy and quality of life among developed and developing countries (98). The authors attribute the outcomes to variation of the health literacy tool among the studies; therefore, more studies are needed to robust evidence to support the relationship. Nevertheless, this study highlights the implication of low literacy that further intervention is needed to improve their knowledge in selfcare.

### Poverty Attribution

Poverty is not merely a result of mental disease, but it also frequently precedes mental illnesses like depression and anxiety, making it a significant risk factor for mental illness and other negative consequences (99). Bullock and Limbert showed that poor respondents are more likely to endorse structural or external attribution for poverty (100, 101). The mean score (4.0) of poverty attribution to the B40 community agrees with the structural barriers, and respondents who refute poverty as a structural issue showed a reciprocal association for depressive and anxiety symptoms. This is supported by a small scale observational study which indicates that individuals who attribute their poverty to structural or government blame are more likely to experience depression (27). Rozmi et al. postulated that respondents who do not blame the government are more self-sufficient and thus experience better financial well-being (102). Therefore, it is crucial to explore individuals’ perceptions of their low-income status to understand their motivation to get out of poverty (27). Short term stipend from the government such as Bantuan Prihatin Rakyat (BPR) in the form of fast cash may ease their financial stress in the short term, but life skill training is needed to get them out of the poverty cycle.

### Stigma and Professional Help-Seeking

Even though the majority of the respondents have a neutral score for self-stigma, the analysis showed respondents who have higher self-stigma were likely to have anxiety symptoms but not for depression. In contrast, respondents with lower mental health-seeking attitudes predict higher depressive symptoms, not anxiety symptoms.

Based on the body of evidence, stigma is one of the well-established determinants for the help-seeking barrier essential for people from low and high-income countries. Lacking a help-seeking attitude prevents them from getting earlier treatment for severe mental health issues. The negative evaluations on professionals possibly derived from negative past experiences and mistrust of the mental health professionals (103). Data from the Institute’s mental health services policy analysis (104) showed an overall limited mental health workforce in this country. Currently, no counsellors are available to handle mild to moderate mental health cases at the primary care clinic. The existing primary healthcare providers who were not trained in mental health were obligated to manage such cases which were first diagnosed with mental health issues. This may create a spillover on public trust in professional help-seeking whenever a mismanaged case occurs.

### Resilience

Approximately one-third of respondents have low resiliency and higher resilience scores moderate depressive and anxiety symptoms. Substantial empirical studies have supported the inverse resilience scores concerning depression and anxiety (105). A 4 years longitudinal study involving a sample of 314 college students in China supported a reciprocal relationship between resilience and mental ill-being (106). Respondents from this study were vulnerable given their low-income status, and almost half of them reported a history of stressful life events. Building resiliency through health promotion is crucial to shield them from the impact of ill mental well-being due to adversity at the verge of the current pandemic financial crisis.

This is one of the first studies that comprehensively explored the various psychological risk factors during the Covid19 pandemic among the low-income group in LMIC. This is a face-to-face study, thus having the benefits of capturing the responses for the non-technology savvy subjects compared to many other online studies done during the pandemic. It also improves rapport and eases the process of obtaining consent. As this was a cross-sectional study of the temporal causal relationships between the independent variables to depression, anxiety and quality of life could not be assigned, and the majority of the sample were heads of household, male and older age group, the findings may not be generalisable to all lower-income populations.

## Conclusions

Mental health problems compromise the quality of life of the low-income group. The prevalence of depression and anxiety symptoms were higher than studies prior to the pandemic. Apart from the socio-demographic, chronic illnesses and stressful life events factors, this study unveiled the positive psychological factors such as resiliency, stigma and help-seeking for mental health. These outputs provide suitable resources for the subsequent psychosocial intervention development for low-income communities. This is much needed to improve mental health status, empower the low-income group population to thrive during this challenging pandemic period and beyond, and shape a healthier human capital for sustainable development.

## Data Availability

Data cannot be shared publicly because of the vulnerability of the population and sensitivity of the subjects. Data are available from University Malaya Research Ethics Committee (UMREC)(contact via Tel No.: 03-79676289 / 6942), Professor Mohd Azlan Shah Zaidi and Professor Mas Ayu Said for researchers who meet the criteria for access to confidential data.

## Supporting Information

**S1 Table. Sample Size According to Study objectives**

(DOCX)

**S2 Table. Reliability of the study tools**

(DOCX)

**S3 Table. Psychosocial risk factors of the B40 respondents from the Petaling district**

(DOCX)

**S4 Table. Health-related profiles of B40 respondents from the Petaling district**

(DOCX)

**S5 Table. Univariable analysis results tables and multivariable variable selection process**

(DOCX)

## Acknowledgement

We are grateful to Ng Yit Han and Nithiah Thangiah for their contribution in statistical analysis.

## Author Contributions

**Conceptualisation:** Wong Min Fui, Mas Ayu Said, Hazreen Abdul Majid, Tin Tin Su, Tan Maw Pin, Rozmi Ismail

**Data Curation:** Wong Min Fui, Mas Ayu Said, Hazreen Abdul Majid

**Methodology:** Wong Min Fui, Mas Ayu Said, Hazreen Abdul Majid, Rozmi Ismail

**Writing:** Wong Min Fui, Mas Ayu Said, Hazreen Abdul Majid, Rozmi Ismail

**Reviewing and Editing:** Wong Min Fui, Mas Ayu Said, Hazreen Abdul Majid, Rozmi Ismail Tin Tin Su, Tan Maw Pin,

**Funding Acquisition:** Mas Ayu Said, Hazreen Abdul Majid, Tan Maw Pin, Tin Tin Su

